# COVID-19-associated ARDS treated with DEXamethasone (CoDEX): Study design and rationale for a randomized trial

**DOI:** 10.1101/2020.06.24.20139303

**Authors:** Bruno M. Tomazini, Israel S. Maia, Flavia R. Bueno, Maria Vitoria A.O. Silva, Franca Pellison Baldassare, Eduardo Leite V. Costa, Ricardo A.B. Moura, Michele Honorato, André N. Costa, Alexandre B. Cavalcanti, Regis Rosa, Álvaro Avezum, Viviane C. Veiga, Renato D. Lopes, Lucas P. Damiani, Flávia R. Machado, Otavio Berwanger, Luciano C.P. Azevedo, for the COALITION COVID-19 Brazil III Investigators

## Abstract

**OBJECTIVES:** The infection caused by the severe acute respiratory syndrome coronavirus 2 (SARS-CoV2) spreads worldwide and is considered a pandemic. The most common manifestation of SARS-CoV2 infection (Coronavirus disease 2019 - COVID-19) is viral pneumonia with varying degrees of respiratory compromise and up to 40% of hospitalized patients might develop Acute Respiratory Distress Syndrome (ARDS). Several clinical trials evaluated the role of corticosteroids in non-COVID-19 ARDS with conflicting results. We designed a trial to evaluate the effectiveness of early intravenous dexamethasone administration on the number of days alive and free of mechanical ventilation within 28 days after randomization in adult patients with moderate or severe ARDS due to confirmed or probable COVID-19.

**METHODS:** This is a pragmatic, prospective, randomized, stratified, multicenter, open-label, controlled trial including 350 patients with early-onset (less than 48h before randomization) moderate or severe ARDS, defined by the Berlin criteria, due to COVID-19. Eligible patients will be randomly allocated to either standard treatment plus dexamethasone (intervention group) or standard treatment without dexamethasone (control group). Patients in the intervention group will receive dexamethasone 20mg IV once daily for 5 days, followed by dexamethasone 10mg IV once daily for additional 5 days or until Intensive Care Unit (ICU) discharge, whichever occurs first. The primary outcome is ventilator-free days within 28 days after randomization, defined as days alive and free from invasive mechanical ventilation. Secondary outcomes are all-cause mortality rates at day 28, evaluation of the clinical status at day 15 assessed with a 6-level ordinal scale, mechanical ventilation duration from randomization to day 28, Sequential Organ Failure Assessment (SOFA) Score evaluation at 48h, 72h and 7 days and ICU-free days within 28.

**ETHICS AND DISSEMINATION:** This trial was approved by the Brazilian National Committee of Ethics in Research (Comissão Nacional de Ética em Pesquisa - CONEP) and National Health Surveillance Agency (ANVISA). An independent data monitoring committee will perform interim analyses and evaluate adverse events throughout the trial. Results will be submitted for publication after enrolment and follow-up are complete.

**ClinicalTrials.gov identifier:** NCT04327401

## INTRODUCTION

In early March 2020, the World Health Organization (WHO) declared the outbreak of a new coronavirus a pandemic^1^. This coronavirus, later named severe acute respiratory syndrome coronavirus 2 (SARS-CoV2), elicited an outburst of severe viral pneumonia (Coronavirus disease 2019 - COVID-19) in mid-December in the Wuhan province, China^2^. The disease spread worldwide, and after three months, countries in all continents, except Antarctica, had registered cases^3^. Considering all cases of COVID-19, estimates suggest that 5% will develop respiratory failure^4^, while in hospitalized patients, up to 40% might develop Acute Respiratory Distress Syndrome (ARDS)^5^, which is the leading cause of death in this population^4^.

Corticosteroids, due to its anti-inflammatory effects^6^, may be a suitable therapy for these patients and have been tested in different scenarios of ARDS^7 8^. A recent trial showed that early use of dexamethasone is safe and reduces the duration of mechanical ventilation in ARDS patients without COVID-19^9^. However, data suggest that corticosteroids use might increase viral load in patients with SARS-CoV-1 infection^10^ and MERS infection^11^, while a meta-analysis showed that corticosteroids are associated with increased mortality in influenza pneumonia^12^. Early use in less severe cases and late use in the course of ARDS might be responsible for the detrimental effects in this population. Current guidelines recommend against using corticosteroids in patients with COVID-19 outside clinical trials^13 14^.

Furthermore, evidence suggests that patients with severe COVID-19 might have a hyperinflammatory state known as cytokine storm. The cytokine profile in these patients resembles the one found in secondary hemophagocytic lymphohistiocytosis (sHLH)^15^, with increased levels of interleukin (IL)-2, IL-6 and tumor necrosis factor alpha. Corticosteroids are one of the therapeutic cornerstones^16^ for treating sHLH.

Therefore, we propose a pragmatic, randomized, open-label, controlled clinical trial, comparing standard treatment versus standard treatment added to early administration of dexamethasone for 10 days in patients with moderate and severe ARDS due to COVID-19. We used the recommendations for Interventional Trials (SPIRIT) guideline for this report^17^, which is presented in the appendix I of the Electronic Supplementary Material (ESM). The steering committee members are shown in the appendix II of the ESM. This manuscript refers to the fifth version of the protocol.

## Methods

### Study design

The COVID-19-associated ARDS treated with DEXamethasone: CoDEX is a pragmatic, prospective, randomized, stratified, multicenter, open-label, superiority, controlled trial including 350 patients with moderate or severe ARDS due to confirmed or probable COVID-19 in 51 Intensive Care Units in Brazil.

We hypothesize that early administration of dexamethasone increases the number of days alive and free of mechanical ventilation in adult patients with moderate or severe ARDS due to SARS-CoV2. The trial is registered with ClinicalTrials.gov (NCT04327401).

### Objectives

Our primary objective is to evaluate the effectiveness of early intravenous (IV) dexamethasone administration on the number of days alive and free of mechanical ventilation within 28 days after randomization in adult patients with moderate or severe ARDS due to confirmed or probable COVID-19. Ventilator-free days (VFD) is defined as being free from invasive mechanical ventilation for at least 48h (successful extubation)^18^. If the patient is re-intubated within 48h of the extubation it will be treated as zero VFD; if re-intubated after 48h, the 48h period will be counted as VFD. Patients discharged from the hospital alive before 28 days are considered alive and free from mechanical ventilation at day 28. Non-survivors at day 28 are considered to have zero VFD.

Secondary objectives are to evaluate the effect of dexamethasone treatment plus standard treatment versus standard treatment alone on the following:

- All-cause mortality rates at 28 days after randomization
- Clinical status of patients at 15 days after randomization using the World Health Organization 6-point Ordinal Scale for clinical improvement (table 1)
- Number of days of mechanical ventilation from randomization to day 28
- ICU free days within 28 days
- Change in the Sequential Organ Failure Assessment (SOFA) Score 48h, 72h and 7 days after randomization

**Table 1.** The 6-point Ordinal Scale

| **Table 1 – The 6-point Ordinal Scale** |  |
| --- | --- |
| 1 | Not hospitalized. |
| 2 | Hospitalized, not requiring supplemental oxygen. |
| 3 | Hospitalized, requiring supplemental oxygen. |
| 4 | Hospitalized, requiring non-invasive ventilation or nasal high-flow oxygen therapy |
| 5 | Hospitalized, requiring invasive mechanical ventilation or ECMO |
| 6 | Death |

### Time schedule and study duration

The planned study duration is five months, being three months of recruitment, one month of follow up and one month for data analysis and manuscript writing. The first patient was enrolled on April 17^th^. The final report and publication are expected to be available on the second half of 2020.

### Eligibility criteria

We will include critically ill patients with ARDS due to confirmed or probable COVID-19 admitted to the ICU. Probable COVID-19 is defined by the presence of symptoms, which are contemplated in the inclusion criteria, travel or residence in a city where community transmission is reported or contact with a confirmed case in the last 14 days prior symptoms onset^19^ and radiological imaging findings compatible with COVID- 19 at the time of inclusion.

After 182 patients have been enrolled, the Steering Committee suggested specific changes on both inclusion and exclusion criteria. The timing of ARDS diagnosis for inclusion changed from 24h to 48h. The rationale for this modification was due to most centers receiving patients intubated in the ICU already with ARDS diagnosis and more than 24 hours of mechanical ventilation, which shortened the time window for recruitment. Additionally, given the widespread use of corticosteroids before ICU admission in Brazil, we allowed inclusion of patients who have previously received one day of corticosteroids during hospital stay, which was not allowed at first. The exclusion criteria were refined by adding three more criteria: use of immunosuppressive drugs, cytotoxic chemotherapy in the past 21 days, and neutropenia due to hematological or solid malignancies with bone marrow invasion.

Each patient must fulfil all the following inclusion criteria to be eligible for enrolment:

- Age ≥18 years old
- Probable or confirmed infection by SARS-CoV2
- Intubated and mechanically ventilated
- Moderate or severe ARDS according to Berlin criteria^20^ (table 2)
- Onset of moderate or severe ARDS in less than 48h before randomization

Exclusion criteria are:

**Table 2.** Berlin Criteria for ARDS diagnosis

| **Table 2 – Berlin Criteria for ARDS diagnosis** |  |
| --- | --- |
| Timing | Within one week of a known clinical insult or new or worsening respiratory symptoms |
| Chest imaging | Bilateral opacities – not fully explained by effusions, lobar/lung collapse, or nodules |
| Origin of edema | Respiratory failure not fully explained by cardiac failure or fluid overload |
| Oxygenation |  |
| Moderate | $100 \text{ mmHg} < \text{PaO}_2/\text{FiO}_2 \leq 200 \text{ mmHg}$ with $\text{PEEP} \geq 5 \text{ cmH}_2\text{O}$ |
| Severe | $\text{PaO}_2/\text{FiO}_2 \leq 100 \text{ mmHg}$ with $\text{PEEP} \geq 5 \text{ cmH}_2\text{O}$ |

- Pregnancy or active lactation
- Known history of dexamethasone allergy
- Daily use of corticosteroids in the past 15 days
- Indication for corticosteroids use for other clinical conditions (e.g refractory septic shock)
- Patients who did use corticosteroids during hospital stay for periods equal or greater than two days
- Use of immunosuppressive drugs
- Cytotoxic chemotherapy in the past 21 days
- Neutropenia due to hematological or solid malignancies with bone marrow invasion
- Patient is expected to die in the next 24 hours
- Consent refusal for participating in the trial

### Study Protocol

#### Randomization and Allocation Concealment

Patients are eligible for enrolment if all the inclusion criteria and none of the exclusion criteria are met. Patients are being randomized in a 1:1 ratio to one of the two groups (figure 1): standard treatment plus dexamethasone (intervention group) and standard treatment without dexamethasone (control group). The randomization list is generated by an independent statistician in random blocks of 2 and 4 in order to preserve the allocation concealment and is stratified by center. Randomization is performed by an online web-based central, available 24h a day. The group treatment is disclosed to the investigator only after all information regarding patient enrolment is recorded in the online system. Patients are screened for enrolment by the principal investigator and the research team at each study center.

### Blinding

This is an open-label trial where the investigators, caregivers and patients are not blinded regarding the intervention. All statistical analyses will be performed in a blinded manner with respect to group allocation.

### Trial intervention and treatment strategy

Each ICU enrolling patients in the trial are encouraged to follow the best practice guidelines and their institutional protocol for the care of critically ill patients with COVID-19. Laboratory testing, hemodynamic management, ventilatory strategy, antibiotics usage, venous thromboembolism and stress ulcer prophylaxis, along with all other ICU interventions are left at the discretion of the ICU team for both intervention and control group.

Patients in the intervention group are receiving after randomization dexamethasone 20mg intravenously once daily for 5 days, followed by dexamethasone 10mg IV once daily for additional 5 days or until ICU discharge, whichever occurs first. Patients in the control group are not receiving dexamethasone.

Although we are not controlling the ventilatory strategy in both groups, physicians are encouraged to comply with the following ventilator strategy: tidal volume (Vt) of 4-6 ml/kg of predicted body weight (PBW), a plateau pressure < 30 cmH_2_0, driving pressure < 15 cmH_2_0, respiratory rate to maintain arterial pH > 7.2 and FiO_2_ and PEEP to keep SpO_2_ ≥ 88% or PaO_2_ ≥ 55 mmHg. Sedation drugs and the use of other strategies for ARDS management such as use of neuromuscular blocking agents, prone positioning, nitric oxide and Extracorporeal membrane oxygenation (ECMO) are left to physicians’ discretion and are registered daily on the study’s electronic case report form (eCRF). Each center is encouraged to follow institutional guidelines for liberation of mechanical ventilation. The study timeline is shown in figure 2.

### Procedure to COVID-19 diagnosis

Due to the possibility of false negative tests, especially on the first days of symptoms^21^, associated with different sensitivities depending of the site of collection, patients with negative laboratory tests included in the study are evaluated by a blinded committee formed by two critical care physicians of the research group with experience treating COVID-19 patients (Adjudication Committee). This committee will take into account the timing of testing, clinical symptoms and analysis of chest image (computed tomography scan of the lungs, or chest X-ray) to define if the patients has COVID-19 infection with negative laboratory tests (probable COVID-19 infection) or if the patient possibly has not COVID-19 infection. Patients with positive polymerase chain reaction (PCR) tests for SARS-CoV2 are deemed to have confirmed COVID-19 infection.

The main analysis will be based on the intention-to-treat principle, with additional sensitivity analysis regarding the COVID-19 infection status (confirmed vs not confirmed).

### Adverse events

The most common adverse effects of corticosteroids use are hyperglycemia and possible increase in infections rates. Data on glycemic control is collected daily until day 14 and data on the development of new infections is collected daily until day 28. For any other adverse events a specific form is available on the eCRF and the data is sent in real time to the coordinating center.

### Handling of protocol deviations

Adherence to protocol and corticosteroids use in both groups is accessed daily. The use of corticosteroids in the control group is not forbidden since critically ill patients might have another indication for corticosteroid use during their ICU stay. However, any use of corticosteroids for treating ARDS and or refractory hypoxemia in the control group is considered protocol deviation. Changes in dosage of dexamethasone or early interruption in the intervention group will also be considered protocol deviation.

If during the trial the patient is deemed to not have COVID-19 infection, which is defined by a negative laboratory tests along with negative evaluation by the Adjudication Committee, the study drug will be stopped, data on these patients will be collected until day 28 and will be included in the final analysis. However, giving the epidemiological context and inclusion criteria, it is expected that only a minority of patients will be in this group.

All centers will receive an initial training session before initiating recruitment to ensure consistency of the study procedures and data collection.

### Data collection and management

Unidentified patient data will be collected through an electronic online data capture tool (REDCap)^22 23^. Demographic and baseline data, height, Simplified Acute Physiology Score (SAPS) 3, use of corticosteroids prior the randomization, and the HScore (table 3) for diagnosis of secondary hemophagocytic lymphohistiocytosis are collected for all patients. The SOFA score is collected on days 1, 2, 3 and 7. Data from gas exchange, lung mechanics, hemodynamic, laboratory data, use of neuromuscular blocking agents, prone positioning and use of extracorporeal membrane oxygenation (ECMO) are collected prior randomization and until day 14.

**Table 3.**
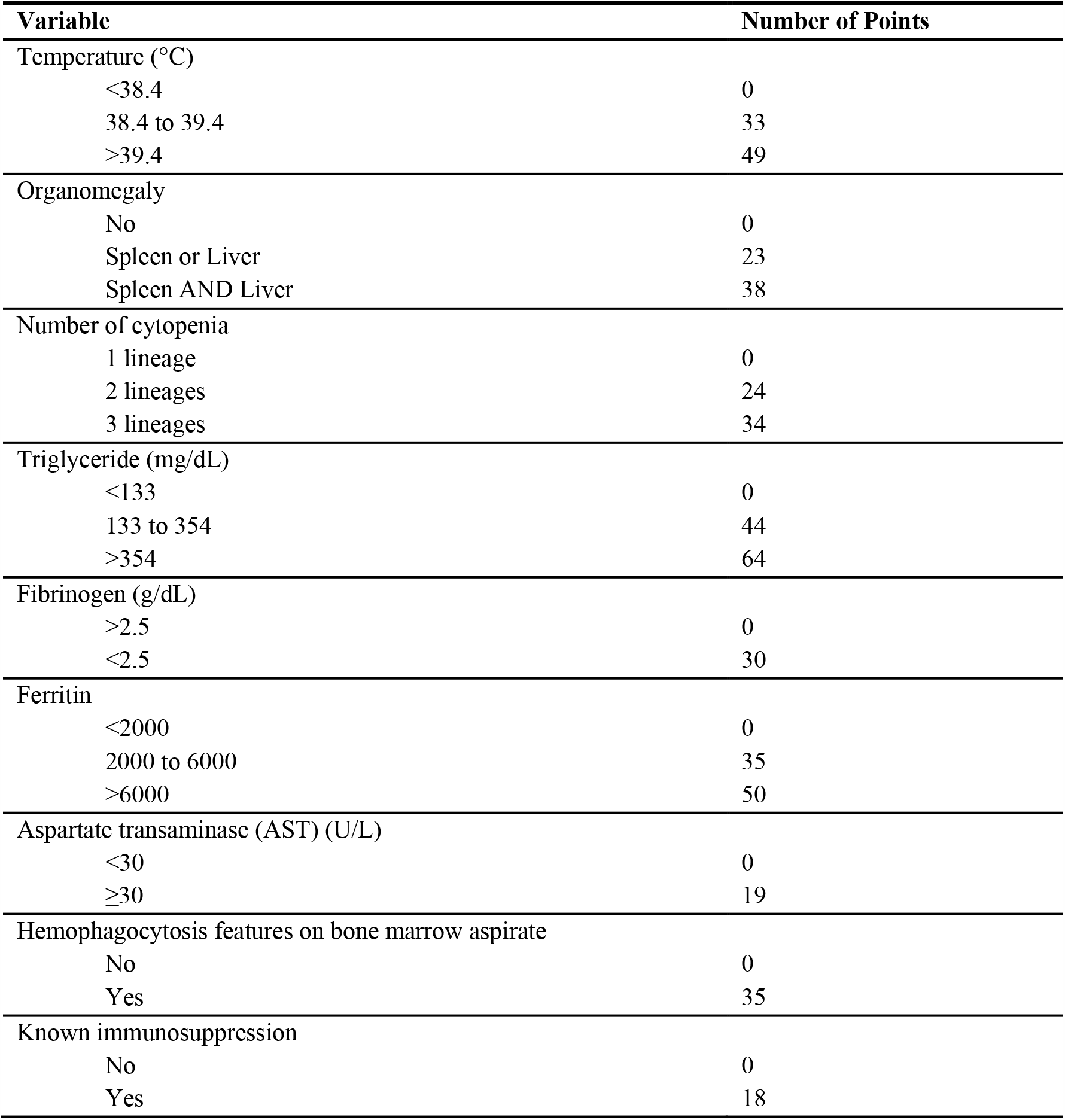
HScore for diagnosis of secondary hemophagocytic lymphohistiocytosis

The use of mechanical ventilation or any other ventilation/oxygen support (high flow nasal cannula, non-invasive ventilation, use of supplementary oxygen) and data on the 6-point Ordinal Scale (table 1) are collected daily until day 28 or until hospital discharge whichever comes first. Vital status at ICU and hospital discharge and any other relevant clinical data such as nosocomial infections, insulin use for glycemic control, antibiotic use, and other therapies for COVID-19 (hydroxychloroquine, chloroquine and azithromycin) are collected.

Data on mechanical ventilation are collected in specific forms with data on date and time of initiation and discontinuation of therapy.

All data are collected through the eCRF and periodically data quality checks will be performed by the Trial Management Committee. Database lock will be carried out after the 28-day outcome is obtained for all patients. Database access will be granted only to steering committee members and statisticians before the main results are published. We plan to share data with other ongoing clinical trials on the same topic for individual patient’s metanalysis. We plan to upload the study dataset to a public database 3 months after database lock.

### Statistical Analysis

#### Sample size calculation

There is a lack of reliable data available in patients with ARDS due to COVID-19 to allow an accurate sample size calculation. We therefore used data from a randomized controlled trial in non-COVID-19 ARDS patients^24^, a well-designed multicenter trial that is representative of ARDS outcomes in Brazil, to calculate the sample size. We assumed a mean of VFD at 28 days of 8 days ± 9 days (standard deviation) in the control group. With a two-sided type I error of 0.05 and power of 80% to identify a difference in three days free of mechanical ventilation between groups, a sample size of 290 patients would be needed. However, in the end of May 2020, before the first interim analysis, after discussing the protocol with the Data Monitoring Committee (DMC), the Steering Committee decided to increase the sample size based on the following rational: Given the uncertainty regarding the normality of distribution of VFD, based on the Pitman Asymptotic Relative Efficiency^25^, the sample size should be increased by 15% to preserve study power coupled with a 4% increase considering possible lost to follow-up and withdrawal of consent. Therefore, a final sample size of 350 patients is needed.

Also, due to the lack of data about ventilator free days in COVID-19 patients, the sample size will be updated using the pooled standard deviation of ventilator free days of the first interim analysis, unless by the time of the first interim analysis all patients have been recruited.

The minimal clinically important difference of three days for VFD was chosen based on other trials^26 27^ along with what is perceived as a significant improvement to the in-hospital complications, costs, and intensive care unit availability, especially in countries with limited resources.

### Interim analysis and safety

Two interim analyses are planned for safety and efficacy evaluation, after 96 patients and 234 patients with the complete follow up to the primary outcome. Based on the results of these interim analyses, the DMC will decide if there is proof beyond a reasonable doubt that the intervention is effective or not safe in this population. The stopping rule for safety will be a p-value <0.01 and for efficacy p-value <0.001 (Haybittle– Peto boundary). The Haybittle–Peto boundary is a conservative stopping rule at interim analysis that has minimal impact in increasing type I error in two-arm trials^28^. The interim analyses will be performed by an external and independent DMC.

### Statistical methods

Main analyses will follow the intention-to-treat principle. For the primary outcome, a generalized linear model will be built with beta-binomial distribution or zero/one inflated beta distribution, with center as random effect and adjusted for age, corticosteroid use before randomization and PaO_2_/FiO_2_ ratio. The effect size will be estimated as mean difference, the respective 95% confidence interval, and hypothesis testing. Missing data on the primary outcome will be dealt with using multiple imputation techniques.

For details regarding the analysis of the secondary outcomes and other analysis, please refer to the Statistical Analysis Plan (SAP) on the appendix III of the ESM.

The significance level for all analyses will be 0.05. There will be no adjustment for multiple testing. All analyses will be performed using the R software^29^ (R Core Team, Vienna, Austria, 2020).

### Subgroup and Sensitivity Analyses

We plan to perform subgroup analysis for the primary endpoint, including interaction parameters in the main model to:

- Age (<60 and ≥60)
- PaO_2_/FiO_2_ ratio (≤100 and >100)
- SAPS3 (<50 and ≥50);
- Duration of symptoms at randomization, days (≤7 and >7)
- Duration of moderate / severe ARDS to randomization, hours (≤24h and >24h to 48h)
- Position at randomization; (prone or supine)
- HScore^30^ (≥169 and <169)
- Use of corticosteroids before randomization
- Use of vasopressors at randomization

We will perform the following pre-specified sensitivity analysis: patients with laboratory confirmed COVID-19, patients with laboratory confirmed and probable COVID-19, per-protocol analysis (patients that received the proposed treatment in the intervention group and patients that not received corticosteroids in the control group) and an as-treated analysis (considering patients which received any dose of corticosteroids in the control group).

### Future additions

We plan to collect blood samples for transcriptomic studies after randomization and after 10 days to evaluate if the treatment effect of dexamethasone changes based on the genetic expression of leukocytes and to follow patients for a period of 12 months in order to perform future analyses on clinical outcomes and quality of life.

### Trial organization

The Steering Committee is constituted by the study investigators of the Coalition COVID-19 Brazil III and will be responsible for the development of the study protocol, manuscript drafts and study submission to publication. All other study committees will report to the Steering Committee.

The Trial Management Committee (TMC) is formed by members of the Coalition COVID-19 Brazil III and is responsible for:

i. Conduction of the study: creating the electronic case report forms (eCRF), designing the investigator manual and the operations manuals, managing and controlling data quality.
ii. Research center management: selecting and training the research centers, assisting the center in regulatory issues, monitoring recruitment rates, monitoring follow-up, sending study materials to research centers.
iii. Statistical analysis and reporting: completing the statistical analyses and helping to write the final manuscript.

The Adjudication Committee is responsible for evaluating all laboratory negative cases of COVID-19. Based on the epidemiology, clinical findings and radiological imaging, the committee will classify patients as probable or negative cases of COVID-19.

The Data Monitoring Committee is composed by an external statistician and experts in critical care medicine independent of the study’s investigators (see ESM Appendix IV for further information). The DMC will be responsible for the interim analysis and will provide guidance to the Steering Committee regarding the continuation and safety of the trial after the interim analyses based on the evidence of significant differences between intervention or control group regarding ventilator free days at day 28, mortality or adverse events.

### Ethical considerations and dissemination

The trial was designed according to the guidelines for good clinical practice and followed the principles of the Declaration of Helsinki and was approved by the Brazilian National Committee of Ethics in Research (Comissão Nacional de Ética em Pesquisa - CONEP). All protocol amendments must be approved by CONEP before its implementation.

Given the growing number of COVID-19 cases in Brazil, most hospitals have adopted total restriction policies in ICU visitation in order to contain viral spreading. Also, we expect that virtually none of the patients will be able to give consent due to their clinical condition. Therefore, the CONEP allowed for different approaches in obtaining the consent from the patients’ legal representatives, such as consent by email or any other digital format and by voice or video. Patients will be included in the study only after the Informed Consent Form is obtained by the study’s investigators. Patients and their legal representatives can withdrawal from the study at any time and for any reason. Patients’ next of kin are assured that this withdrawal will not have any impact regarding the patients’ care. Before withdrawal patients or their legal representative will be asked if data can continue to be collected, despite receiving the study interventions. Patients who withdrawal consent from the study will not be replaced by other participants.

The study will be submitted for publication after completion irrespective of its findings. The manuscript elaboration will be an inalienable responsibility of the Steering Committee. The main paper will be authored by the steering committee members plus the principal investigators of the 10-top enrolling sites, which can contribute intellectually to the manuscript.

## Discussion

This is the first randomized controlled trial evaluating the efficacy of early dexamethasone administration in moderate and severe ARDS caused by the SARS-CoV2 virus. Corticosteroids have been used in ARDS treatment for almost 50 years^31^. However, there is still controversy around the efficacy of this treatment. The literature suggests a potential benefit of early administration in more severe cases with a possible influence on the outcome depending on the ARDS cause (bacterial vs. viral pneumonia, primary vs. secondary ARDS). Also, most of the published data is from small, retrospective studies in heterogeneous populations.

The most common adverse effects of corticosteroids use is hyperglycemia, but as shown in a recent trial^9^, patients receiving dexamethasone had a similar frequency of hyperglycemia (76%) as compared to controls (70%). Also, the trial showed no difference in new infections in the ICU between groups.

Our trial has significant strengths compared to the published literature. The study population will be homogenous comprising only critically ill patients with moderate or severe ARDS. We offer a precise and reproducible intervention protocol and we will include patients in the early phase of ARDS. Early ARDS phase probably coincides with a later phase in the disease process, which might reduce the risk of increased viral replication induced by the study drug as suggested by previous authors for MERS virus^11^ and SARS- CoV1 infection^10^.

We acknowledge our trial has some limitations. It is an open-label trial, which can interfere in the use of other immunomodulatory therapies such as the use of convalescent plasma, tocilizumab or hydroxychloroquine, especially in the control group. However, we choose an objective primary outcome with clear definitions, which reduces the influence of the open label nature in the outcome assessment.

If we confirm our hypothesis of benefit of using dexamethasone in ARDS due to SARS-CoV2 infection, the consequences for public health will be enormous, especially considering the COVID-19 pandemic. Given the unprecedented impact in global health and the lack of ICU beds in most countries during the pandemic, an increase in days alive and free of mechanical ventilation should help unburden the health care systems worldwide and will represent a noteworthy improvement in ARDS treatment.

## Data Availability

This is a protocol for an ongoing randomized controlled trial. We plan to upload the study dataset to a public database 3 months after database lock.

